# Eco-anxiety among children and young people: A systematic review of social, political, and geographic determinants

**DOI:** 10.1101/2023.12.19.23300198

**Authors:** Claire L Niedzwiedz, Shamal M Kankawale, S Vittal Katikireddi

**Affiliations:** School of Health and Wellbeing, University of Glasgow, Clarice Pears Building, 90 Byres Road, Glasgow, G12 8TB

## Abstract

Eco-anxiety refers to distress arising from environmental degradation, especially concerning climate change, with children and young people (CYP) likely to be particularly susceptible. This systematic review examined social, political, and geographic factors associated with eco-anxiety among CYP. We searched 11 databases, Google Scholar and pre-print servers up to August 2024, and citations to January 2025. Quality was assessed using the Mixed Methods Appraisal Tool (MMAT). 69 studies (42 quantitative, 16 qualitative, 11 mixed-methods) were included in the narrative synthesis. Most studies used non-probability sampling, limiting generalisability, and were from the Global North. Findings were grouped into three categories of determinants: social (age, gender, media exposure, socioeconomic and educational contexts, intergenerational relations, peer and cultural norms), political (distrust, government inaction, individual views, and participation), and geographic (exposure to environmental hazards, cross-country differences, urban or rural residence). Further study is needed to understand how eco-anxiety develops and varies globally, particularly regarding how social, political and geographic factors intersect, and its impacts on mental health and wellbeing.

---

Climate change is leading to a range of cumulative, interconnected and escalating effects on global health.^1,2^ The associated health impacts include shifts in the incidence and distribution of vector-borne diseases, heat-related morbidity and mortality linked to extreme weather, amongst others^3^. Climate change disproportionately affects children and young people (CYP), who are projected to experience between two and seven times more extreme climatic events over their lifetimes, compared to older generations^4^. These effects are expected to be most severe in the Global South^4^. In particular, under Paris Agreement commitments, a child born in 2020 is estimated to face seven times the risk of extreme heatwaves and twice the risk of wildfire exposure compared to a person born in 1960, and this risk is even higher for children living in poverty and high climate risk regions^4–6^. These stressors are likely to have severe consequences for physical health, but also mental health and wellbeing^6^. Despite this, children’s mental health is almost invisible within national adaptation policies on climate change^7^.

Climate change impacts mental health via direct, indirect and overarching pathways^8,9^. Direct effects arise from exposure to extreme weather events, such as heatwaves^10^, and slower-onset changes, such as drought.^11^ Indirect effects may occur via disruptions to social, economic and other determinants of mental health, such as food insecurity, displacement and interruptions to education^8^. Additionally, the anticipation, awareness and understanding of climate change can also affect mental wellbeing, even among people without firsthand experience of climate-related events^12^. This overarching psychological burden may particularly affect CYP, who are increasingly exposed to educational content and media coverage concerning climate change and the lack of sufficient policy action^13,14^. Adolescence, a developmental period marked by heightened interest in global and environmental issues,^13^ also coincides with the onset of many common mental health disorders.^15^ Therefore it is essential to support CYP in developing adaptive coping strategies to manage the psychological impacts of climate awareness and related environmental concerns.

A range of terms – such as climate worry, eco-anxiety, climate anxiety, solastalgia and ecological grief – have emerged to describe the psychological responses to climate and environmental changes, predominantly within Western contexts^9,14,16^. Worry, reflects apprehensive thoughts and emotional discomfort related to perceived future threats or anticipated negative outcomes^17,18^. Although often regarded as a negative cognitive-emotional state, worry can serve an adaptive function by motivating individuals to take preparatory or protective action^19,20^. In relation to climate change, climate worry is typically characterised by verbal-linguistic processing (rather than visual imagery) focused on current or potential changes in the climate system and the anticipated consequences^9,21^. Importantly, climate worry has been related to increased engagement in pro-environmental behaviours^22,23^.

However, when worry becomes persistent, difficult to control and repetitive, it may signal the presence of anxiety^17,21^. Eco-anxiety encompasses a broader range of distressing emotions linked to environmental degradation, including – but not limited to – climate change, and reflects concern over the numerous and intersecting threats posed by the ecological crisis^24^. Climate anxiety, considered a subset of eco-anxiety, specifically denotes psychological distress related to the climate crisis^9^.

Several psychometric instruments have been developed to assess levels of eco- and climate anxiety, including the Climate Change Anxiety Scale (CCAS)^16^ and Hogg Eco-Anxiety Scale (HEAS)^25^. The CCAS comprises four dimensions: cognitive-emotional impairment (e.g. experiencing nightmares about climate change), functional impairment (e.g. concerns about climate change that interfere with daily responsibilities - such as work or school), behavioural engagement (e.g. taking actions like recycling or consuming less meat), and personal experience (e.g. being directly affected by climate-related events)^16^. The HEAS also comprises four dimensions - affective symptoms, rumination, behavioural symptoms and anxiety related to personal impacts - but adopts a broader scope by incorporating concerns about a wider range of environmental issues beyond climate change, such as species extinction, pollution and deforestation^25^. High or chronic levels of eco-anxiety could lead to emotional fatigue or psychological burnout, affecting capacity to engage constructively with environmental challenges or participate in collective climate action^26^, however research is still in its infancy. A systematic review of 12 studies examining the health implications of eco-anxiety found the evidence base to be of limited methodological quality, but associations with functional impairment, symptoms of depression, anxiety, post-traumatic stress disorder (PTSD), stress, insomnia, and poorer self-rated mental health were suggested.^27^ A meta-analysis of 25 studies assessing the relationship between climate anxiety (restricted to studies using the CCAS) and wellbeing also demonstrated a moderate negative correlation, that may be stronger among people with a stronger environmental identity^28^. Climate worry has also been associated with future risk of clinical levels of anxiety among a large sample of adults across Europe^29^. Identifying and understanding the diverse factors that contribute to eco-anxiety is essential to formulate comprehensive strategies to support psychological wellbeing and encourage engagement in constructive environmental action.

The emergence and intensity of eco-anxiety is likely to be shaped by a complex interplay of individual, social, political and environmental factors^9,30^. A social-ecological approach can help to understand the various factors that may contribute to eco-anxiety among CYP^30^. Disparities in vulnerability to climate change and differential exposure to environmental threats across sociodemographic groups may contribute to unequal experiences of eco-anxiety. Disadvantaged communities, particularly those residing in regions disproportionately affected by environmental hazards, often experience poorer mental health outcomes - especially in the aftermath of extreme weather events^31,32^ - due to limited adaptive resources and heightened exposure to adverse social and economic determinants of health.

However, little research has examined whether these groups also experience correspondingly high levels of eco-anxiety. Indeed, critiques of the concept highlight the individualised and depoliticised mainstream approach to eco-anxiety, suggesting it represents a “privileged anxiety”^33,34^, dominated by white experiences^35^, mainly from the Global North^9^.

Although eco-anxiety has attracted increasing global attention across media discourse^36^, clinical settings^37,38^, and academic research^28,39,40^, there remains a notable gap in understanding the various social, political, and geographic determinants that influence its development and persistence among CYP. Existing literature reviews have focused on defining and measuring eco-anxiety among CYP^41^, primarily within the Global North setting, or exploring its psychological impacts^13,42^, with relatively limited investigation into the broader contextual determinants shaping its expression in younger populations^30^. This review therefore seeks to address this gap by synthesising evidence from a wide range of studies to provide a comprehensive overview of the factors influencing eco-anxiety experiences among CYP.

## Methods

The study used a systematic review methodology and took a narrative synthesis approach to analyse literature focused on the determinants of eco-anxiety among CYP. The review is reported according to the Preferred Reporting Items for Systematic Reviews and Meta-Analysis (PRISMA) guidelines^43^. The review design was guided by the PECO framework: **P**opulation included children and young people aged under 25 years; **E**xposure related to social, political, and geographic determinants; **C**omparator included different levels of the determinant being studied (e.g. for gender: young woman, man, or other gender); **O**utcome was eco-anxiety. The systematic review was registered on the PROSPERO database (ID=CRD42023440162, updated in October 2024 to broaden searches and inclusion criteria).

### Eligibility criteria

Studies were eligible for inclusion if they investigated eco-anxiety among children and/or young people, defined as aged under 25 years, consistent with World Health Organization and United Nations classifcations^44^. Studies in which the majority of participants were aged under 25 years (or the median age of participants was <25 years), or where subgroup analyses allowed for extraction of data specific to this age group, were also considered for inclusion. All empirical study designs were considered, including quantitative, qualitative, mixed-method studies, and systematic reviews and meta-analyses, provided they included human participants.

To be included, studies had to contain information on the potential social, political or geographic determinants of eco-anxiety. This encompassed quantitative studies that assessed associations between specific variables (e.g. gender, political orientation, or country of residence) and eco-anxiety, as well as qualitative studies where such factors emerged as relevant themes. Studies that assessed the level or prevalence of eco-anxiety in a particular country or group, or which validated an appropriate measurement tool were also considered for eligibility. Outcome measures included eco- or climate anxiety, operationalised as anxiety, worry, fear, or psychological distress in relation to environmental issues or climate change. Studies conducted in any geographic location or setting were eligible, but only English language studies were included.

Articles were excluded if they did not report original research (e.g. narrative reviews, opinion pieces, or commentaries), did not include human participants, or examined populations predominantly aged 25 years and over without providing disaggregated data for CYP. Studies assessing general mental health outcomes (e.g. generalized anxiety or depressive symptoms) in relation to extreme weather or environmental changes without consideration of eco-anxiety were also excluded. Studies published in a language other than English were not considered for inclusion.

### Search strategy

A systematic search of electronic databases was conducted during August 2024. A librarian at the University of Glasgow was consulted for advice on the search strategy. Searches (title and abstracts) were performed using EBSCOhost which included APA PsycArticles, APA PsycInfo, Child Development & Adolescent Studies, CINAHL, EconLit, GreenFILE, Health Source: Nursing/Academic Edition, Psychology and Behavioral Sciences Collection, and SocINDEX. Additional searches were conducted via Ovid MEDLINE (no limiters) and Web of Science (title and abstracts). Google Scholar (first 10 pages of results) was also used to search for relevant grey literature (e.g. reports). Relevant unpublished academic articles were searched for using pre-print servers (MedRxiv and PsyArxiv). Backward and forward citation searches were also performed to ensure completeness in the search process up to January 2025. An example of the search performed in MEDLINE is included in Appendix 1. The search terms were as follows: (“eco-anxiety”, “ecoanxiety”, “climate anxiety”, “climate change anxiety”, “environmental anxiety”, “fear of climate change”, “eco-distress”, “ecological stress”, “climate-related stress”, “climate distress”, “climate worry”, “climate concern”, “environmental worry”, “environmental concern”, “environmental distress”, “climate emotions”, “ecological emotions”) AND (“child”, “children”, “youth”, “young people”, “teenagers”, “teens”, “adolescence”, “juvenile”, “youngster”, “adolescent”, “minor”, “kid”). Searches were not limited by date of publication or language.

### Study selection

Search results were exported to Covidence and duplicate articles removed. Zotero was used to manage references. Articles were first screened from titles and abstracts within Covidence, using the inclusion and exclusion criteria to eliminate irrelevant articles. The remaining articles were screened based on their full texts and reasons for exclusion were recorded. Two reviewers (CLN and SVK) independently conducted title and abstract screening, followed by full-text screening. Conflicts were resolved through discussion.

### Data extraction

Study characteristics (authors, title, publication year, country, location/setting, sample size, sampling strategy, dates of data collection), study design (e.g. quantitative, qualitative, mixed-method, systematic review, meta-analysis, observational or experimental, cross-sectional or longitudinal), participant characteristics (e.g. age, gender, ethnicity, socioeconomic position), social determinants of eco-anxiety (e.g. gender, education level, income), geographic determinants of eco-anxiety (e.g. country, exposure to climate-related events), political determinants of eco-anxiety (e.g. voting preference), outcome (e.g. eco-anxiety, climate anxiety, climate worry), outcome measurement tool used (CCAS, HEAS), analysis approach, key findings and limitations were extracted from each included study. Under key findings we also extracted summary measures for eco-anxiety if the study reported a mean, percentage, or similar summary statistic. We used the Artificial Intelligence (AI) tool Elicit (elicit.com) to facilitate data extraction^45,46^, the results of which were exported into Excel and thoroughly checked by CLN, and further data extraction was performed manually by CLN where required (e.g. from supplementary material). Studies which were unable to be extracted via Elicit (N=1^47^) were manually extracted by CLN. SVK completed data extraction for a random 10% sample and all data extracted were further checked by SMK.

### Quality appraisal

Given the diverse range of study designs included, the Mixed Methods Appraisal Tool (MMAT)^48,49^ was selected as the most appropriate quality assessment instrument. The MMAT assesses the quality of qualitative, quantitative, and mixed-method studies, focusing on five core methodological quality criteria. For example, qualitative studies are assessed using the following questions: 1) Is the qualitative approach appropriate to answer the research question?; 2) Are the qualitative data collection methods adequate to address the research question?; 3) Are the findings adequately derived from the data?; 4) Is the interpretation of results sufficiently substantiated by data?; 5) Is there coherence between qualitative data sources, collection, analysis and interpretation?^48^. To provide a crude overall quality score, ‘yes’ answers were scored as 1, and ‘no’ or ‘can’t tell’ were scored as 0. The scores for each item were summed to provide an overall quality score ranging from 0 to 5, with those scoring 3 or more classified as moderate to high quality. Quality appraisal was conducted manually by CLN (checked by SMK), recorded in an Excel spreadsheet and a random 10% sample was independently completed by SVK. Alongside the quality appraisal, the type of article was also noted (e.g. peer-reviewed journal article, dissertation, pre-print, conference abstract).

### Data synthesis

The data synthesis involved an iterative process of summarising and analysing the quantitative and qualitative findings from included studies using the data extraction spreadsheets, re-reading the articles, and noting key themes, statistics and quotations. A narrative synthesis approach was employed to interpret and integrate the evidence on the factors associated with eco-anxiety. Relevant guidelines for narrative synthesis were consulted to improve transparency^50^. The narrative synthesis involved identifying common themes, patterns, and trends across studies. These themes were categorised based on social, political and geographic determinants, allowing for the organisation and comparison of findings. Further subgroup themes were identified within the overall themes, some of which were pre-defined based on previous literature – for example, age and gender. Others emerged from the synthesis, for example, if more than one study highlighted the theme. The limitations of the included studies and heterogeneity in the findings were noted, paying attention to contradictory results. Synthesised findings were presented in a narrative format, supported by appropriate tables and quotations. The authors confirm that all data generated or analysed during this study are included in this published article.

## Results

### Study selection

The database searches yielded a total of 11,782 results. 99 articles were also identified from other sources (e.g. citations and grey literature searches). After removing 1309 duplicates, 10,572 titles and abstracts were screened for inclusion. Following title and abstract screening, 217 articles were screened using the full texts. Ultimately, 69 met the eligibility criteria after excluding 148 full texts (Fig. 1 provides full details of the search process and reasons for exclusion).

**Figure 1:**
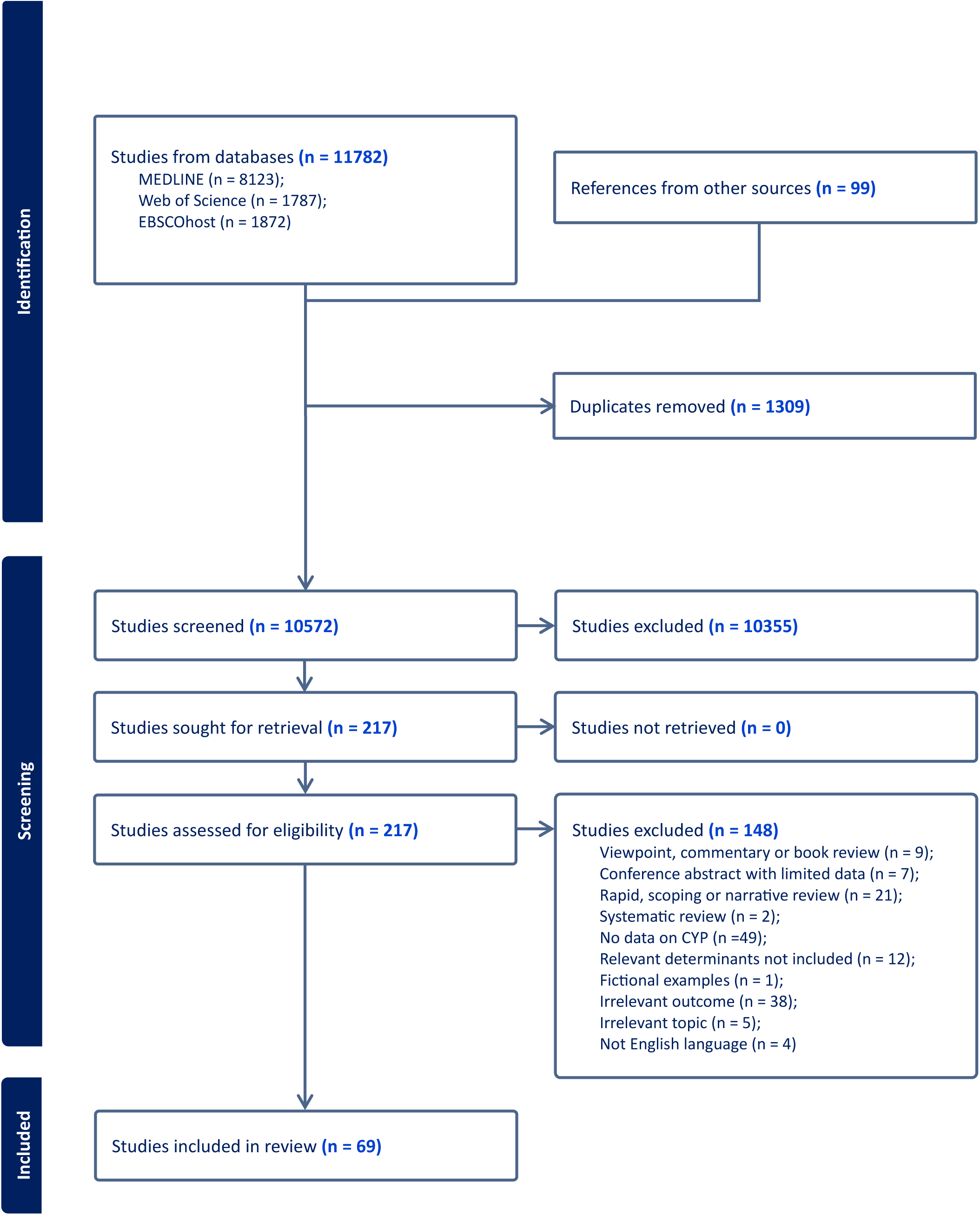
PRISMA flowchart

### Study characteristics

Sixty-nine articles were included in the systematic review (summarised in Table 1). All selected studies were published between 1995 and 2024, with a notable increase from around 2021. There were 42 quantitative, 16 qualitative and 11 mixed-method studies. Of the quantitative studies, most were cross-sectional, with only two studies identified as longitudinal^51,52^. The sample size included within studies varied from seven participants in a qualitative study examining eco-anxiety experiences and coping strategies^53^, to 139,941 in a cross-sectional study of Norwegian school pupils^54^. In terms of geographic location, there was a higher representation of countries from the Global North like Australia, the United Kingdom, Canada, Sweden and the USA compared to those from the Global South. Only two studies that focused on a single country were identified from Africa, these were from Tanzania^55^ and Kenya^56^. Only one study was identified concentrating on a country from South East Asia, which was based within the Philippines^57^, and very few were from East Asia (China^58^), and South America (Brazil^59^). Amongst the cross-national studies included, again the Global South was underrepresented, with the USA and European countries, particularly the United Kingdom represented in the greatest number of studies^20,60–62^.

**Table 1:** Summary of included studies.

Study quality was variable, with the MMAT checklist score ranging from 0 to 5, with an average of 3 (full results of the quality appraisal can be found in Appendix 2). Across all studies, most adopted convenience, snowball or quota sampling, but a few quantitative studies included samples which were believed to be nationally representative of the general population, based on probability sampling^20,54,63^. The absence of a standard measure of eco-anxiety led to a variety of measurement approaches being implemented within quantitative studies. In studies that used a validated scale, the Hogg Eco-Anxiety Scale^25^, Climate Change Anxiety Scale^16^, Climate Change Worry Scale^21^ and Reser’s (2012) Climate Distress Scale^64^ were used. In other studies, which tended to be older studies published before validated measures were developed, the authors created their own questions^47,65^, which were often simple single-item measures. A notable trend was the use of online methods for data collection, both for online surveys and qualitative interviews, perhaps reflecting that data collection for many recent studies tended to take place during or after the COVID-19 pandemic period. Most quantitative studies involved survey data collection, with a few analysing secondary data of an existing dataset^20,51,55,66^. A couple of studies included an experimental component which tested the effect of media exposure on eco-anxiety^58,67^. A notable limitation across all studies was the lack of ethnic diversity and inclusion of Indigenous groups^60,68–70^, as well as the over- representation of people from more advantaged socioeconomic backgrounds^59,69^ and young women^67,71–76^.

Amongst the qualitative and mixed-method studies, data were collected via focus groups^59,76–79^, in-depth interviews^53,68,70,72,78,80–88^, open-ended survey questions^69,71,75,89,90^, participatory action research^91,92^, auto-photography^72^, diary^93,94^, drawing^81,94^, participant observation^94^ and Q sorts^78^ methods. Particular limitations concerning the mixed-method studies were the small sample size of the quantitative components^69,85^, and the lack of analysis of the divergencies and inconsistencies between the quantitative and qualitative findings. The qualitative studies included also sometimes lacked detail on their analysis approaches^68,83,91^.

## Synthesis of findings

The analysis identified several themes within the three categories of determinants contributing to experiences of eco-anxiety (Table 2). Social determinants included factors such as age, cohort and developmental stage, gender, media exposure, intergenerational relations, peer and cultural norms, and socioeconomic and educational contexts. Political determinants encompassed government and institutional inaction, distrust and feeling of betrayal, and political views, action and participation. Geographic determinants captured the influence of place-based experiences, including direct exposure to environmental hazards, cross-country differences, and urban–rural distinctions. Each overarching set of determinants is discussed in detail below (further details in Appendix 3).

**Table 2:**
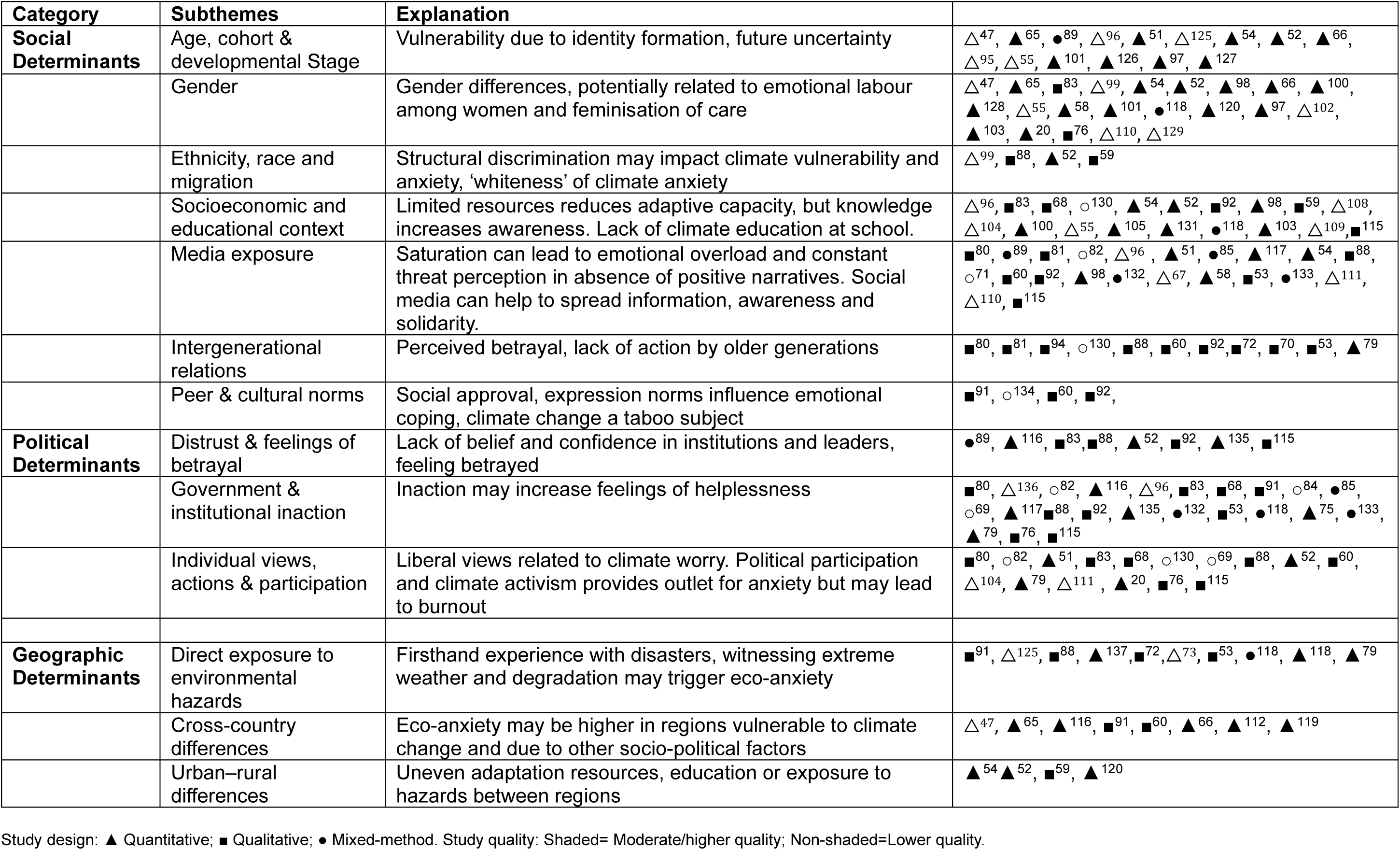
Subthemes within the categories of social, political and geographic determinants of eco-anxiety.

### Social determinants

#### Age, cohort and developmental stage

The findings from the reviewed studies underscored age and developmental stage as key determinants of eco-anxiety. A consistent pattern emerged across multiple studies showing that adolescents and young adults reported heightened levels of climate-related worry, eco-anxiety, and distress^95^. Several studies documented that late adolescents and young adults often engage more deeply with climate issues, with this engagement intensifying as their cognitive and emotional capacities mature^65,80,89^. This pattern was reinforced within longitudinal studies, which demonstrated an age-related increase in climate change worry from 15 to 21 years, indicating that as individuals aged, their perception of environmental threats became more acute^52^. The eight-year longitudinal study conducted by Sciberras and Fernando (2022) found that as age increased from 10-11 years up to 18-19 years increasing levels of climate change-related worry were observed^51^. Similarly, in their longitudinal study, Prati et al (2022) observed that climate change worry increased from age 15 years to around 23-25 years, after which a small decrease was observed^52^. Metsäranta (2021) found eco-anxiety was experienced more strongly among people aged 24-29 years compared to those aged 15-23 years in a sample of Finnish young people^96^. Similar findings were also observed by Prencipe et al (2023), who found older youth in Tanzania reported more climate change distress^55^, and by Donati et al (2024) who found climate worry was higher among older adolescents than younger adolescents^97^.

However, a cross-national study across 23 countries mostly in Europe found little difference in climate worry between age groups^20^, but differences between countries were not explored. In a Norwegian study of people aged 13-19 years, those aged 15-16 years were more likely to be worried about climate change, but the relationship with age was non-linear, decreasing among those aged 18-19 years^54^. In a mixed-methods study focusing on Swedish children and young adults, Ojala (2021) noted climate worry was stronger among adolescents (aged around 16-17 years) and young adults (aged around 20-25 years), compared to children (aged around 11-12 years)^89^. In a qualitative study by Chou et al (2023), the authors noted that children aged around 11-12 years may experience more climate-related distress compared to younger children as they develop and become more able to envisage a hypothetical future^59^. These results suggest that eco-anxiety may not be uniformly distributed across age groups, but may instead be shaped by psychological development, making adolescence a particularly critical period.

#### Gender

Consistent gender differences in the experience of eco-anxiety were found, with most studies suggesting that young women and girls express higher eco-anxiety levels compared to other genders^16,20,54–56,65,97–103^. Leonhardt et al. (2022) found that adolescent girls were more likely to experience eco-anxiety than boys: 14% of girls reported being very worried about climate change, compared to 7% of boys, and 28% of boys reported being not worried at all, compared to 10% of girls, in their study including 128,484 participants^54^. In their logistic regression models adjusted for sociodemographic factors and leisure activities, girls had 2.60 (95% CI: 2.53–2.67) higher odds of eco-anxiety compared to boys, and this association persisted after further adjustment for mental health and health behaviours. In a study including 2,652 high school pupils in Kenya, young women were more likely to report being afraid of climate change compared to men (42.3% compared to 33.8%)^56^. This gender disparity was also found within several other studies, with a few exceptions.

Comparing climate activists with non-climate activists in Turkey, Ediz and Yanik (2023) found no notable differences in climate change anxiety between genders^104^. Hill-Harding et al. (2024) found no apparent differences between genders for most results in their study of students at a large UK university^75^. However, overall findings suggest gender differences in eco-anxiety emerge early and generally persist across developmental stages and different country contexts. Parsons et al. (2024) found that even among a relatively gender progressive country, young women in New Zealand often felt greater sense of responsibility for pro-climate action compared to men, which is linked to the general feminisation of care practices.

> *“is the emotional and psycho-social burden[s] of caring [for the environment] and [concern about] climate change falls to a huge extent onto women… [This results in] negative impacts [for me and] a lot of women’s psyches, like carrying this burden. I think about [how I can] help fix [the climate challenges faced by] underprivileged women in other countries as well. (Izadora, FG1)”, p1451*^76^.

#### Ethnicity, race and migration

Very few studies reported on ethnic differences in eco-anxiety and studies generally lacked adequate representation of ethnic minority groups. One study found that students from ethnic minority groups reported lower levels of eco-anxiety compared to those from the white majority group in a study within English secondary schools^99^. A Swedish study found no difference in climate change worry when comparing people from Swedish and foreign national backgrounds^100^. A number of studies included people from Māori and other ethnic minority groups in Aotearoa (New Zealand), suggesting these group may be particularly affected by eco-anxiety^68,69,88^.

#### Socioeconomic context

Several studies highlighted the socioeconomic and educational contexts as potential influential factors for eco-anxiety, but findings differed depending on the aspect of socioeconomic position studied (e.g. education level, occupation, or income) and scale (e.g. individual, parental, household, or area-level). According to Leonhardt et al. (2022), adolescents who perceived their family’s financial situation as good had lower risk of being worried about climate change, compared to those who perceived their financial situation as poor, in the large Norwegian study^54^. They also found that the level of parental education was consistently associated with eco-anxiety, compared to those with no higher education, young people whose parents had higher education had 1.57 (95 CI: 1.52–1.61) higher odds of being worried about climate change^54^, which was also found in several other studies, including within Sweden^100^, Portugal^105^ and Tanzania^55^. In a study based in Australia, those living in more disadvantaged areas (according to the Index of Relative Socio-economic Disadvantage) had higher odds of experiencing eco-anxiety compared to those living in more advantaged areas^95^. Among UK residents, higher socioeconomic status (measured by the Family Affluence Scale^106^) was associated with higher levels of eco-anxiety^90^, however, another study found that young adults in an unfavourable financial situation had higher odds of experiencing stress related to the climate crisis in Poland^103^.

Chou et al. (2023) elaborated in their qualitative study using focus groups with participants aged 5-18 years in three areas of Brazil^59^, that the socioeconomic context shapes awareness and engagement with climate change:

> *‘‘There are two types of rich and poor people who deal with this situation: the rich ones either don’t care, they only think about money – or are informed and try to do as much as possible, because they have the money for it. There are two types of poor people, who either have no place to find information – or they have information, but they don’t have money to afford organic food (12,F)”,* p260^59^.

Further, they found that the groups who were more aware and engaged with climate change issues belonged to the wealthier social classes and experienced more eco-anxiety^59^. This group comprised children and adolescents whose parents were more engaged in environmental action, or who attended private schools which had climate change integrated into their curricula. The influence of parental occupation was also noted by a participant in another qualitative study, who noted that their mother was a journalist and felt well-informed by them^59^. Working in extreme temperatures was also linked with climate change distress in a Tanzanian study^55^, but there was a general lack of studies examining occupational influences. Another study highlighted how youth climate activists from privileged backgrounds had greater access to emotional and social resources to navigate eco-anxiety, further highlighting inequality in resilience and response capacity^68^. Other studies noted the educational context and that students enrolled in environment-related courses tended to report higher levels of eco-anxiety^83,107–109^. However, these cross-sectional studies do not imply a causal relationship since students likely self-select based on their concern about the environment.

#### Media exposure

Media exposure emerged in several studies as a potential determinant of eco-anxiety^58,67,71,81,82,85,96,107,110,111^. In multiple qualitative studies, increased exposure to climate-related news, especially through social and mass media, was mentioned as a key theme. Studies highlighted that media allow the distribution of helpful information which can inform people about environmental issues^80^, but it can also intensify feelings of helplessness, especially when messaging is fear-based or lacks hopeful framing^111,112^.The frequency of media use and attention given to climate change news were key predictors of climate anxiety in a study based in the USA, with media exposure variables explaining around a third of the variance in climate anxiety scores^111^.

Many studies mentioned social media as an important factor affecting how children and young people experience eco-anxiety^51,54,71,85^. A mixed-method study by Gunasiri et al. (2022^b^) including 46 participants aged 18-24 years in Australia found that negative stories about climate change shared on social media can contribute to feelings of hopelessness, guilt, shame and anxiety, which can be overwhelming^85^.

> *“I do get a lot of climate anxiety when I read the stuff about how June was the hottest month, I’m like yeah I can’t do anything about that. [Interview participant 7]”,* p6^85^.

On the other hand, social media was also reported to have a positive role, providing a platform for young people’s voices and demonstrating action:

> *“You can be speaking to people on the other side of the globe that you’d never interact with at any other point in your life and you’re able to build a community and a network… “, p7*^85^.

In another Australian study by Sciberras and Fernando (2022), greater societal engagement, including consumption of news relating to international affairs, was related to high and increasing levels of climate-related worry over time^51^. A study across eight countries also found that participants reported increased climate anxiety due to social media coverage^71^. In contrast, the large cross-sectional Norwegian study by Leonhardt et al. (2022) found that individuals who expressed higher levels of concern about climate change tended to spend less time using social media^54^. Despite some variation, the overall pattern suggests that media can act as both a conduit for climate awareness and action, as well as a vector for psychological burden.

#### Intergenerational relations

A few studies indicated that intergenerational relations, including trust in older generations and worry about future generations, are potentially related to eco-anxiety^53,72,77,79,108^. Boyd et al. (2024) highlighted a ‘generational misalignment’, where youth report frustration and distress arising from perceived climate inaction by older generations^79^. This lack of trust in adults’ willingness or ability to mitigate climate threats contributed to feelings of betrayal and powerlessness among young people with pre-existing mental health problems in Australia^79^. Frustration at the burden of trying to fix previous generations’ actions was similarly highlighted in another qualitative study^70,77^.

> *“Yeah, I’m also angry at previous generation because actually, it’s us, it’s us who have to live with this. . . Like people who are 70 will die in like 30 or 20 years even, you know. . . it’s me who is 11 years old that will live in 40 years, 50 years, 60 years. . . Probably I’m going to live that again and again because others who are 70 years old today will have passed away when I’ll be 70 years old. . . But I will be there, I will see everything that degrades.”– Participant 15, 11 years old,* P10^70^.

Smith et al. (2023) also documented how young women expressed eco-anxiety in the context of contemplating future family planning, citing concerns about the environmental legacy being inherited from previous generations^72^. These findings suggested that diminished intergenerational trust may exacerbate eco-anxiety, but also influence life choices and outlooks.^108^

> *“I mainly think about my own children that I want to have in the future […] And, you know, since I feel like they will be close to me because they are my family, I feel like I’m already worried about them. And these children don’t even exist and this future that doesn’t even exist. But it’s something that worries me the most. I think about my future family living in a world with climate change.” (Participant 7, female, 23)”, P34*^53^

#### Peer and cultural norms

The influence of peer and cultural norms emerged as correlates of eco-anxiety in some studies^47,51,65,77^. Thomas et al. (2022) found that young people often described their climate-related emotions as shaped not only by personal experiences, but by the social environments in which they are embedded, including peer interactions and broader cultural narratives^77^. Cultural expectations regarding environmental responsibility, particularly in collectivist or activist-oriented communities, may heighten pressure to respond emotionally or behaviourally to the climate crisis. Sciberras and Fernando (2021) reported that individual worry profiles may be influenced by shared social and cultural contexts, where normative beliefs about climate change can either validate or suppress eco-anxiety^51^. In a qualitative study using participatory action methods, young people also highlighted the stigma surrounding climate-related concern and the unwillingness to talk about these issues in countries like Jamacia (where some older people considered extreme weather to be a normal occurrence), but also in the UK (where it has been considered a taboo subject)^91^. They also mentioned struggling with whether to discuss their concerns with family, friends and colleagues. A culture of denial was also a found to be a source of eco-anxiety in a small mixed-method study among adolescents participating in environmental groups at the University of Vermont, Canada^82^. Being ‘shamed’ by peers, being belittled and experiencing derision related to their interest in climate issues was highlighted by several participants in a small study based within a Welsh school:

> *“I was made fun of and called Greta Thunberg for speaking out loud for defending my opinion. I know for a fact that I am not the first one, and sadly will not be the last one to experience it… I believe that is the school’s duty to stop shaming people for their beliefs and start to give better education. (Sara, YP5, 2166-2174)”,* P80^113^.

### Political determinants

#### Distrust

Studies consistently revealed concern from CYP regarding their lack of inclusion and representation in decision-making processes and their lack of trust in government and institutions, which can exacerbate eco-anxiety^62,69,87,90,91,114^. The cross-sectional study by Hickman et al. (2021) with data from 10,000 young people across ten countries found that participants felt more betrayal (highest in Brazil, India and the Philippines) than reassurance towards the government and that children believed government responses to the climate crisis have been inadequate, which correlated with climate anxiety and distress^62^. Barnes (2022) found that participants expressed pervasive distrust in government actors, associating this with heightened existential concern and hopelessness about the future^83^. Similar sentiments were reported by Thomas et al. (2022), where US participants described disappointment and anger toward political leaders as intensifying their eco-anxiety^77^ and in a study focused on Australia exploring CYP’s emotional responses to climate change.

> *“We have a few more Independent politicians in Parliament now […] so I think that is a positive… I feel more hopeful, but the trust is still low due to the previous government when nothing happened. (Young person, age 24).”, P9*^115^

Other studies further elaborated that CYP often perceive political leaders as unresponsive or indifferent to the urgency of the climate crisis, resulting in frustration, alienation, and a diminished sense of agency^59,77,87,91,92^. These studies suggest that eco-anxiety is not merely a reaction to environmental degradation, but also a response to political systems perceived as inadequate or disingenuous in addressing the crisis.

#### Government and institutional inaction

Government and institutional inaction was identified as a recurring theme contributing to eco-anxiety. The large cross-sectional study by Hickman et al. (2021), highlighted that young people’s negative perceptions of governmental responses to climate change were associated with increased levels of distress^116^. Moreover, government inaction was frequently mentioned to be intertwined with political interests and industry lobbying, exemplified by Myers et al. (2022) in an Australian study, which highlighted young people’s frustration and anger as they witnessed fossil fuel industries influencing policymaking and distributing misinformation^88^. This hindered effective climate action leading to reduced trust and contributing to eco-anxiety, especially when young people felt their concerns were being dismissed. These findings were corroborated in another study by Boyd et al. (2024)

> *“They’re really letting us down, like, [the Prime Minister] is putting so much money towards fossil fuels. When we really need to focus on the environment at the moment, because what’s the point of making money if you’re not going to have a planet? You know those politicians who are meant to oversee the stuff, they’re in their like, 60s, 70s. So by the time it actually hits really hard, a lot of them will probably not be here anymore. So it’s a bit discouraging as well. Like, it’s sort of been relegated to us. (Client8).”, P1027*^79^

Several studies revealed that perceived governmental inaction, lack of transparency, and symbolic rather than substantive climate policies erode public confidence and contribute to emotional distress^53,59,68,72,79,82–85,91,95,117^. Hill-Harding et al. (2024) examined students’ emotional responses to climate change among university students in the UK and also found that perceptions of inadequate institutional responses, in this case the university, can exacerbate eco-anxiety^75^.

#### Political views, participation and actions

The literature consistently highlighted other political factors may shape eco-anxiety, including political views and participation. Parsons et al. (2024) found that youth engaged in climate activism frequently expressed both empowerment and psychological strain, highlighting the emotional weight of taking on political responsibility when institutional responses are seen as inadequate^76^. Sciberras and Fernando (2022) found that adolescents with consistently high or increasing climate change-related worry were more politically engaged^51^. Gunasiri et al. (2022) similarly found that participants who took climate action reported higher levels of worry, but that there were also positive benefits (e.g. feeling more in control)^85^ and in other studies the development of a social network which validated their identity was highlighted^80^. Some studies also noted concern around the potential repercussions of protest involvement^88^. Other studies also noted a more liberal or left-leaning political orientation was associated with higher eco-anxiety among CYP^83,111^.

### Geographic determinants

#### Direct exposure to environmental hazards

The direct experience of climate-related events (e.g. wildfire) was highlighted in several studies that explored CYP’s personal experiences^57,59,73,85,88,91,118^. A cross-sectional study by Vercammen et al. (2023) in the USA reported that individuals who had direct experience of climate change had higher mean scores for climate distress and eco-anxiety compared to those who had not encountered such impacts, even when taking into account age, gender, education level, family affluence, urban/rural residence and ethnicity^118^. The cross-sectional study conducted by Lykins et al. (2023) investigated Australian youth mental health in the aftermath of the Black Summer Bushfires during 2019/20, highlighting the mental health impact of localised events related to climate change on the individuals affected directly^73^. Young people directly exposed to the bushfires experienced higher levels of climate change distress and concern compared to those who were not directly exposed. The study found that the proximity of the bushfire event, whether in terms of physical distance, social, or temporal aspects, did not appear to have an impact on anxiety levels^73^.

Simon et al. (2023) similarly found that young people living in the Philippines, where many people constantly experience first-hand the effects of climate change-related typhoons and droughts, were prone to experiencing eco-anxiety, but this also motivated them to take climate action^57^.

Qualitative research in New Zealand highlighted the potential impact of living in a coastal community, where one participant saw first-hand the erosion of the foreshore on his journey to school^88^. Chou et al. (2023) also highlighted that having family members impacted by climate-related events, such as flooding, led to feelings of fear amongst some participants in a qualitative study based in Brazil^59^. People living in low-lying countries, such as the Philippines and Jamacia, reported experiencing despair that their homelands may cease to exist in the future due to sea-level rise and flooding^91^. Hearing the rain was also highlighted as a trigger for eco-anxiety among those affected by flooding in an Australian study focusing on young people with mental health problems^79^.

> *“And at night time that rain, for me, it was triggering. It was like, ‘Are we going to be flooded’? You know, I’m safe, but it still doesn’t help you not think about it. Think about others and yeah, animals, wildlife, all those things that are put out of place. I think that affects a lot of people too. (Client7).” P1028* ^79^

#### Cross-country differences

The study by Hickman et al. (2021) suggested that the climate vulnerability of regions may be an important geographic determinant of eco-anxiety, with notable cross-country differeces^116^. Surveying 10,000 CYP across ten countries—including both high-risk and less-affected regions—the study found that youth living in areas more vulnerable to climate impacts (e.g. Philippines and India) reported higher levels of climate-related worry and distress^62^, compared to countries such as Finland and France. In another multi-country study, Lau et al. (2024) explored differences in emotional engagement with climate change including climate anxiety, finding that China had the highest levels, compared to Portugal, South Africa, United Kingdom and the USA^61^. Other cross-national studies found few differences between levels of climate worry between countries^65,119^. Differences in measurement and scoring approaches used across quantitative studies made it difficult to synthesise results from single country studies, therefore we focused on the multi-country studies (see Appendix 4 for full table of results). However, these are also limited by the lack of nationally representative samples.

#### Urban-rural differences

Potential differences in the manifestation of eco-anxiety were found based on the urban or rural residence of the individual. For example, Strife et al. (2012) conducted interviews with urban American children who expressed heightened environmental anxiety, influenced by their exposure to urban environmental degradation^81^. In contrast, studies such Ndetei et al. (2024) suggested that youth living in rural or semi-urban regions may experience eco-anxiety differently, with their concerns more closely tied to direct interactions with the natural environment and localised climate impacts^56,104^. Young people living in rural areas were found to experience a higher level of climate worry compared to those living in urban areas of Kenya^56^. Chou et al. (2023) also identified regional disparities in Brazilian children’s climate awareness, emphasising that urban access to information and activism differed from rural lived experiences^59^. Adolescents living in urban areas of Norway were more prone to eco-anxiety, compared to those living in rural areas^54^. Similarly, Prati et al. (2022) found living in the countryside to be negatively correlated with climate change worry^52^.

## Discussion

This systematic review synthesised 69 studies exploring the determinants of eco-anxiety in children and young people. While we found a range of factors to be related to eco-anxiety, including social (age and developmental stage, gender, socioeconomic context, media exposure, peer and cultural norms, and intergenerational relations), political (distrust, government inaction, and individual views and participation), and geographic (direct experience of environmental hazards, cross-country differences, and urban-rural status), the review also identifies a number of significant gaps in the literature and the overall lack of methodological quality affecting the evidence base.

The review highlighted the importance of age, with eco-anxiety generally increasing throughout adolescence and peaking in early adulthood^52^. This trajectory appears to reflect developmental changes in cognitive, emotional, and moral reasoning. Further exploration of the longitudinal development of eco-anxiety throughout the life course is needed to fully understand age and developmental trajectories, as well as differentiating these patterns from secular trends in an evolving climate. Gender also emerged as a strong correlate, with young women and girls more likely to report eco-anxiety across a number of diverse contexts. Stigma and societal norms relating to emotional expressiveness and empathy might partly account for women and girls exhibiting a greater tendency for heightened eco-anxiety^59,71^. Socioeconomic context presented a more complex picture, with both higher and lower socioeconomic position linked to increased eco-anxiety, likely through different mechanisms— awareness and exposure, respectively. Political determinants, particularly government inaction, were consistently reported as correlates of climate distress^116^. Many studies highlighted how a lack of trust in leadership and intergenerational inequity contributes to perceptions of helplessness, betrayal, and existential worry. At the same time, youth agency and activism emerged as both a response to eco-anxiety and a coping strategy, though one that carries emotional burdens.

Geographically, the experience of eco-anxiety seems heightened in climate-vulnerable regions and among populations directly exposed to environmental hazards, such as wildfires^73,116^ and flooding^79^. Urban–rural differences were observed, but varied by country^52,54,120^.

There was a general lack of diversity in the countries included, with several studies from Australia, Europe and the USA, but very few including populations living in Africa, Asia and South America. Further research is needed from countries in the Global South to understand the applicability of eco-anxiety among different cultures and to fully explore differences between countries and cultures. There was an important gap concerning research which linked aspects of the environment, such as air pollution, flood risk, and temperature, to experiences of eco-anxiety. While this review covered a diverse range of determinants, evidence on specific factors, such as ethnicity and migration status, were lacking, limiting the depth of synthesis for some aspects. It should also be highlighted that studies sometimes mentioned other factors which were not fully investigated, such as religiosity, spirituality^84^, and sexual orientation^103^. Most studies also recruited participants using online methods, which likely excludes those without internet access, such as people who live in more remote areas and Indigenous groups who may be at higher risk from climate change impacts. Studies frequently used non-probability sampling, limiting generalisability and the ability to adequately compare differences in prevalence between countries. Mixed-method studies were of generally of poorer quality, particularly relating to their quantitative components. A key gap is the lack of studies which took an intersectional approach, there is therefore a need to examine the potentially compounded risks experienced by those with multiple disadvantages.

This systematic review has several strengths that build on existing reviews^13,41,42,121^. The comprehensive search process covered several interdisciplinary databases and included relevant studies across various contexts and populations. Including qualitative, quantitative, and mixed-method studies further enriched the depth of the review. Moreover, we assessed the quality of studies using the MMAT toolkit, but there may be some limitations with this approach and the quality appraisal process is inherently subjective^48^. However, most previous reviews have not assessed the quality of existing studies^13,42,121^. We included only studies that were published in English, which may have introduced bias, potentially omitting valuable insights from non-English language literature. Heterogeneity in the definition and measurement of eco-anxiety limited the ability to conduct meta-analysis and made comparisons between studies challenging. There is generally a lack of conceptual clarity around eco-anxiety and related terms, therefore there is a chance we may have overlooked studies that used other similar terms. Furthermore, our interest was in exploring the determinants of eco-anxiety, rather than attempting to establish causal relationships. This reflects the relatively early development of the evidence base, the need to understand who is most affected and to identify factors that warrant further investigation. Finally, while efforts were made to identify grey literature, language and publication biases may have influenced the pool of included studies. The narrative synthesis is also somewhat influenced by the authors’ biases and subjective interpretations.

A significant research gap pertains to the long-term impacts of eco-anxiety on the mental health and wellbeing of CYP. There is limited research on how early-life eco-anxiety influences later psychological trajectories, coping strategies, and decision-making behaviours over time. There is a need for longitudinal studies that track eco-anxiety during youth into adulthood, currently limited by the lack of available data and measurement tools. The limited exploration of effective coping mechanisms for CYP to manage their eco-anxiety, may hinder the development of strategies to mitigate its potential adverse impacts. Furthermore, research is needed to explore the influence of ethnicity, race and migration status and its intersection with other factors such as gender, education and income. Studies that explore environmental and geographic factors, like living in urban, rural or coastal settings, are also needed to understand eco-anxiety experiences in different places.

Our review underscores the need for climate adaptation and mitigation policies that centre the experiences and future of CYP. Young people increasingly report feeling like their views are not being heard and that they lack a say in important decisions which will affect them the most.^116^ This can foster feelings of disempowerment and hopelessness. Political leaders can take important steps towards more inclusive and equitable decision making. Reform of voting systems which often exclude young people is one way in which CYP could feel more empowered^122,123^. Teachers, health professionals and parents/carers, amongst others, could more actively encourage civic engagement among young people, whilst also taking effective action themselves^124^.

To conclude, this systematic review demonstrates that eco-anxiety among CYP is likely shaped by a range of social, political, and geographic determinants. While experiences of eco-anxiety vary across contexts, certain groups—particularly adolescents, young women, individuals living in climate-vulnerable regions, and those exposed to government inaction or intense negative media coverage—may be particularly affected. The review highlights the need for consistent, validated measures to assess eco-anxiety, as well as greater representation of populations within the Global South and generally more survey data which are nationally representative. It also underscores the importance of examining eco-anxiety not only as an individual psychological response, but as a reflection of broader systemic failures, including political inaction, social inequality, and environmental injustice. Managing eco-anxiety likely requires a dual approach: enhancing individual coping and resilience through education, support, and open dialogue, while simultaneously advancing structural changes that restore trust, promote intergenerational equity, and meaningful involvement of CYP in political decisions. As the climate crisis continues to evolve, understanding and addressing the emotional dimensions of its impact especially for younger generations must be a key priority for researchers, policymakers, educators, and mental health practitioners.

## Supporting information

PRISMA checklist

## Data availability

This is a systematic review study based on findings in published literature and did not involve analysis of newly generated data. All data extracted are available in the online appendix.

## Code Availability

This systematic review did not generate any new code.

