## appendix 4 for "Eco-anxiety among children and young people: A systematic review of social, political, and geographic determinants"

| **First author** | **Lau** | | | | **Rizeq** | | **Szagun** | | **Hokka** | **Hickman** | **Lau** |
| --- | --- | --- | --- | --- | --- | --- | --- | --- | --- | --- | --- |
| **Measure (range)** | **Emotional engagement with climate change (0-68)** | | **Anxious about climate change (1-5)** | | **Level of worry about climate change (1-6)** | | **Strength of anxiety about future environmental destruction (1-6)** | | **Very worried about greenhouse effect** | **Extremely worried about climate change** | **Very much anxious about climate change** |
|  | **Mean** | **SD** | **Mean** | **SD** | **Mean** | **SD** | **Mean** | **SD** | **%** | **%** | **%** |
| **Country** |  |  |  |  |  |  |  |  |  |  |  |
| Australia |  |  |  |  |  |  |  |  |  | 25.0 |  |
| Brazil |  |  |  |  |  |  |  |  |  | 29.0 |  |
| China | 31.6 | 12.8 | 2.1 | 1.2 |  |  |  |  |  |  | 12.0 |
| Estonia |  |  |  |  |  |  |  |  | 28.0 |  |  |
| Finland |  |  |  |  |  |  |  |  | 32.0 | 18.0 |  |
| France |  |  |  |  |  |  |  |  |  | 18.0 |  |
| Germany |  |  |  |  |  |  | 5.1 | 0.8 |  |  |  |
| India |  |  |  |  |  |  |  |  |  | 35.0 |  |
| Nigeria |  |  |  |  |  |  |  |  |  | 22.0 |  |
| Philippines |  |  |  |  |  |  |  |  |  | 49.0 |  |
| Portugal | 29.7 | 13.4 | 1.7 | 1.3 |  |  |  |  |  | 30.0 | 10.8 |
| Qatar |  |  |  |  | 3.8 | 1.2 |  |  |  |  |  |
| Russia |  |  |  |  |  |  | 5.1 | 0.7 | 28.0 |  |  |
| South Africa | 27.8 | 15.2 | 1.6 | 1.3 | 3.8 | 1.3 |  |  |  |  | 9.5 |
| Turkey | 31.6 | 12.8 |  |  |  |  |  |  |  |  |  |
| UK | 26.9 | 14.5 | 1.8 | 1.3 |  |  |  |  |  | 20.0 | 9.8 |
| USA |  |  | 1.8 | 1.3 |  |  |  |  |  | 19.0 | 11.0 |

Appendix 4: Summary of results from cross-national studies
