## appendix 1 for "Eco-anxiety among children and young people: A systematic review of social, political, and geographic determinants"

Ovid MEDLINE(R) ALL <1946 to August 02, 2024>

1	Child/ or Adolescent/ or Minors/ or Schools.mp. or Schools, Nursery/	3331770

2	(child or child$).mp.	2861399

3	(schoolchild or schoolchild$ or (school adj child) or (school adj child$)).mp.	42172

4	(kid or kids).mp.	10917

5	(toddler or toddler$).mp.	15738

6	adolesc$.mp.	2364079

7	(teen or teens or teenager or teenager$).mp.	29776

8	(boy or boy$).mp.	183412

9	(girl or girl$).mp.	180453

10	(minors or minors$).mp.	8398

11	juvenil$.mp.	110997

12	youth$.mp.	114337

13	kindergar$.mp.	9009

14	(schools or preschool$ or (pre adj school$)).mp.	1180737

15	((primary adj school$) or (secondary adj school$)).mp.	31488

16	((elementary adj school) or (elementary adj school$)).mp.	12782

17	((high adj school$) or highschool$).mp.	43368

18	(schoolage or (school adj age$) or schoolage$).mp.	30510

19	exp anxiety/	119682

20	Stress, Psychological/	136837

21	exp panic/ or exp fear/	41653

22	(worr$ adj2 ((climate$ or environment$ or eco$) not economic$)).mp. [mp=title, book title, abstract, original title, name of substance word, subject heading word, floating sub-heading word, keyword heading word, organism supplementary concept word, protocol supplementary concept word, rare disease supplementary concept word, unique identifier, synonyms, population supplementary concept word, anatomy supplementary concept word]	148

23	(fear adj2 ((climate$ or environment$ or eco$) not economic$)).mp. [mp=title, book title, abstract, original title, name of substance word, subject heading word, floating sub-heading word, keyword heading word, organism supplementary concept word, protocol supplementary concept word, rare disease supplementary concept word, unique identifier, synonyms, population supplementary concept word, anatomy supplementary concept word]	285

24	(distress$ adj2 ((climate$ or environment$ or eco$) not economic$)).mp. [mp=title, book title, abstract, original title, name of substance word, subject heading word, floating sub-heading word, keyword heading word, organism supplementary concept word, protocol supplementary concept word, rare disease supplementary concept word, unique identifier, synonyms, population supplementary concept word, anatomy supplementary concept word]	226

25	(concern$ adj2 ((climate$ or environment$ or eco$) not economic$)).mp. [mp=title, book title, abstract, original title, name of substance word, subject heading word, floating sub-heading word, keyword heading word, organism supplementary concept word, protocol supplementary concept word, rare disease supplementary concept word, unique identifier, synonyms, population supplementary concept word, anatomy supplementary concept word]	9397

26	(angst adj2 ((climate$ or environment$ or eco$) not economic$)).mp. [mp=title, book title, abstract, original title, name of substance word, subject heading word, floating sub-heading word, keyword heading word, organism supplementary concept word, protocol supplementary concept word, rare disease supplementary concept word, unique identifier, synonyms, population supplementary concept word, anatomy supplementary concept word]	2

27	(anxi$ adj2 ((climate$ or environment$ or eco$) not economic$)).mp. [mp=title, book title, abstract, original title, name of substance word, subject heading word, floating sub-heading word, keyword heading word, organism supplementary concept word, protocol supplementary concept word, rare disease supplementary concept word, unique identifier, synonyms, population supplementary concept word, anatomy supplementary concept word]	518

28	("climate change" or "change, climate" or "changes, climate" or "climate" or "climate changes" or "climates" or "effect, greenhouse" or "environment" or "environmental impact" or "environmental impacts" or "environmental pollution" or "environments" or "greenhouse effect" or "impact, environmental" or "impacts, environmental" or "pollution, environmental").mp.	1171591

29	exp "Climate Change"/ or exp "Climate"/ or exp "Environmental Pollution"/ or exp "Greenhouse Effect"/ or exp "Environment"/	1996696

30	1 or 2 or 3 or 4 or 5 or 6 or 7 or 8 or 9 or 10 or 11 or 12 or 13 or 14 or 15 or 16 or 17 or 18	4316913

31	22 or 23 or 24 or 25 or 26 or 27	10485

32	19 or 20 or 21	274695

33	28 or 29	2794615

34	32 and 33	26351

35	31 or 34	36431

36	30 and 35	8123

37	Child/ or Adolescent/ or Minors/ or Schools.mp. or Schools, Nursery/	3331770

38	(child or child$).mp.	2861399

39	(schoolchild or schoolchild$ or (school adj child) or (school adj child$)).mp.	42172

40	(kid or kids).mp.	10917

41	(toddler or toddler$).mp.	15738

42	adolesc$.mp.	2364079

43	(teen or teens or teenager or teenager$).mp.	29776

44	(boy or boy$).mp.	183412

45	(girl or girl$).mp.	180453

46	(minors or minors$).mp.	8398

47	juvenil$.mp.	110997

48	youth$.mp.	114337

49	kindergar$.mp.	9009

50	(schools or preschool$ or (pre adj school$)).mp.	1180737

51	((primary adj school$) or (secondary adj school$)).mp.	31488

52	((elementary adj school) or (elementary adj school$)).mp.	12782

53	((high adj school$) or highschool$).mp.	43368

54	(schoolage or (school adj age$) or schoolage$).mp.	30510

55	exp anxiety/	119682

56	Stress, Psychological/	136837

57	exp panic/ or exp fear/	41653

58	(worr$ adj2 ((climate$ or environment$ or eco$) not economic$)).mp. [mp=title, book title, abstract, original title, name of substance word, subject heading word, floating sub-heading word, keyword heading word, organism supplementary concept word, protocol supplementary concept word, rare disease supplementary concept word, unique identifier, synonyms, population supplementary concept word, anatomy supplementary concept word]	148

59	(fear adj2 ((climate$ or environment$ or eco$) not economic$)).mp. [mp=title, book title, abstract, original title, name of substance word, subject heading word, floating sub-heading word, keyword heading word, organism supplementary concept word, protocol supplementary concept word, rare disease supplementary concept word, unique identifier, synonyms, population supplementary concept word, anatomy supplementary concept word]	285

60	(distress$ adj2 ((climate$ or environment$ or eco$) not economic$)).mp. [mp=title, book title, abstract, original title, name of substance word, subject heading word, floating sub-heading word, keyword heading word, organism supplementary concept word, protocol supplementary concept word, rare disease supplementary concept word, unique identifier, synonyms, population supplementary concept word, anatomy supplementary concept word]	226

61	(concern$ adj2 ((climate$ or environment$ or eco$) not economic$)).mp. [mp=title, book title, abstract, original title, name of substance word, subject heading word, floating sub-heading word, keyword heading word, organism supplementary concept word, protocol supplementary concept word, rare disease supplementary concept word, unique identifier, synonyms, population supplementary concept word, anatomy supplementary concept word]	9397

62	(angst adj2 ((climate$ or environment$ or eco$) not economic$)).mp. [mp=title, book title, abstract, original title, name of substance word, subject heading word, floating sub-heading word, keyword heading word, organism supplementary concept word, protocol supplementary concept word, rare disease supplementary concept word, unique identifier, synonyms, population supplementary concept word, anatomy supplementary concept word]	2

63	(anxi$ adj2 ((climate$ or environment$ or eco$) not economic$)).mp. [mp=title, book title, abstract, original title, name of substance word, subject heading word, floating sub-heading word, keyword heading word, organism supplementary concept word, protocol supplementary concept word, rare disease supplementary concept word, unique identifier, synonyms, population supplementary concept word, anatomy supplementary concept word]	518

64	("climate change" or "change, climate" or "changes, climate" or "climate" or "climate changes" or "climates" or "effect, greenhouse" or "environment" or "environmental impact" or "environmental impacts" or "environmental pollution" or "environments" or "greenhouse effect" or "impact, environmental" or "impacts, environmental" or "pollution, environmental").mp.	1171591

65	exp "Climate Change"/ or exp "Climate"/ or exp "Environmental Pollution"/ or exp "Greenhouse Effect"/ or exp "Environment"/	1996696

66	37 or 38 or 39 or 40 or 41 or 42 or 43 or 44 or 45 or 46 or 47 or 48 or 49 or 50 or 51 or 52 or 53 or 54	4316913

67	58 or 59 or 60 or 61 or 62 or 63	10485

68	55 or 56 or 57	274695

69	64 or 65	2794615

70	68 and 69	26351

71	67 or 70	36431

72	66 and 71	8123
